# Chloroquine for treatment of COVID-19 - a pig in a poke?

**DOI:** 10.1101/2020.07.06.20147470

**Authors:** R.J. Brüggemann, D.J.A.R. Moes, K.P. van Rhee, N.E. van ’t Veer, B.C.P. Koch, M. van Rossum, A. Vermeulen Windsant - van den Tweel, M.H.E. Reijers, R.R.J. van Kimmenade, J.C. Rahamat- Langedoen, T.C.D. Rettig, R. van Raalte, J. van Paassen, F.N. Polderman, P.D. van der Linden, T. Frenzel, Q. de Mast, D.M. Burger, J. Schouten, F.L. van de Veerdonk, P. Pickkers, R. ter Heine

**Author notes:** Corresponding author: Dr. Roger JM Brüggemann, Radboud university medical center, Department of Pharmacy, 864, POBOX 9101, 6500 HB Nijmegen.

## Abstract

**Objective:** Chloroquine has been frequently administered for treatment of coronavirus disease 2019 but there are serious concerns about its efficacy and cardiac safety. Our objective was to investigate the pharmacokinetics and safety of chloroquine in hospitalized COVID-19 patients.

**Design:** A prospective observational study.

**Setting:** Dutch hospitals

**Patients:** Patients admitted to the hospital for treatment of COVID-19.

**Interventions:** Pharmacokinetic sampling

**Measurements:** The plasma concentrations of chloroquine and desethylchloroquine and QTc time.

**Main Results:** A total of 83 patients were included. The median (IQR) plasma concentration chloroquine during treatment was 1.05 μmol/L (0.63 - 1.55 μmol/L). None of the patients reached exposure exceeding the concentration to inhibit SARS-CoV-2 replication by 90% (_IC90_) of 6.9 μM. Furthermore, ΔQTc >60 milliseconds occurred after initiation of chloroquine treatment in 34% patients and during treatment QTc ≥500 milliseconds was observed in 46% of patients.

**Conclusions:** Recommended dose chloroquine treatment results in plasma concentrations that are unlikely to inhibit viral replication. Furthermore, the incidence of QTc prolongation was high. The preclinical promise of chloroquine as antiviral treatment in patients with COVID-19 is overshadowed by its cardiac toxicity and lack of effective exposure. It is unlikely that a positive clinical effect will be found with chloroquine for treatment of COVID-19.

## Introduction

At the end of 2019, the SARS-CoV-2 virus emerged as a cause of severe respiratory illness beginning in Wuhan, China. Soon thereafter, the World Health Organization declared COVID-19 a pandemic [1]. One study found 90% inhibition of viral replication at a concentration of 6.9 μM (IC_90_), which, according to the authors, was a clinically achievable concentration[2]. Another study found an even higher IC_90_ of chloroquine (>10 μM) against SARS-CoV-2[3]. These preclinical studies have caused the wide scale repurposing of chloroquine as a treatment for COVID-19. At present, it remains unknown whether the preclinical determined *best-case* therapeutic threshold of 6.9 μM is achieved in COVID-19 patients and whether this threshold can be directly translated to the clinic, while there are major concerns about the cardiac safety of chloroquine[4, 5]. We, therefore, investigated the pharmacokinetics and cardiac safety of chloroquine in hospitalized COVID-19 patients.

## Materials and Methods

This was a prospective observational study of hospitalized COVID-19 that were treated with chloroquine from March 20, 2020 until May 1, 2020 in 2 academic hospitals and 3 teaching hospitals in the Netherlands. The institutional review boards of the Radboud University Medical Center and Leiden University Medical Center waived the need for formal ethics approval as all data were collected anonymously using minimally invasive measures. Chloroquine was dosed at a loading dose of 600 mg followed by 300 mg 12 hours later on day 1, followed by 300 mg bidaily for a total treatment duration of five days[6]. Plasma concentrations of chloroquine and its metabolite desethylchloroquine were measured in samples collected as a 4-point pharmacokinetic curve over an 8 hour interval or in randomly collected surplus samples from routine laboratory monitoring by means of a validated bioanalytical assay. Furthermore, chloroquine and metabolite protein unbound concentrations were determined in a subset of 20 random samples using ultrafiltration. Lastly, the unbound fraction of chloroquine and desethylchloroquine were determined at a concentration of 1 and 10 μM in six-fold in the same medium as used for the experiments by Wang et al (Dulbecco’s Modified Eagle Medium with 10% fetal bovine serum). Electrocardiography was performed at baseline and during treatment at the discretion of the treating physician and the QTc-interval was calculated by computerized interpretation. An increase in QTc of >60 ms or a QTc of >500 ms was considered a major risk factor for Torsades de Pointes[7].

## Results

A total of 83 patients were included in this prospective observational study. The patient characteristics are reported in table 1. The patients in our population had a median age of 65 years with an interquartile range (IQR) of 57-73. All but 2 patients were admitted to the intensive care unit and 60% of the patients was male. The majority of the patients concomitantly used other QTc-prolongating medication (see table 1), with propofol as the most frequently used drug in this critically ill a population. The measured concentrations of chloroquine, its active metabolite desethylchloroquine and the individual sum concentration of both chloroquine and its metabolite per day are depicted in figure 1. None of the measured concentrations of chloroquine, nor the sum of chloroquine and its metabolite reached the best-case IC_90_ found *in vitro* for chloroquine. Furthermore, it was found that the median (interquartile range) protein unbound fractions of chloroquine and desethylchloroquine in clinical samples were 18.3% (16.2-22.4%) and 20.2% (18.2-25.2%), respectively. The median unbound fractions of chloroquine and desethylchloroquine in cell medium with 10% fetal bovine serum at the 1 μM concentration level were 62.0% (IQR 58.4-633%) and 71% (IQR 69.1-73.0%), respectively. At the 10 μM concentration level these were 84.1% (IQR 83.8-84.4%) and 88.3% (IQR 87.6-88.7%)

**Table 1:** detailed description patients

| Variable | No (%) of patients (N=83) |
| --- | --- |
| **Demographics** |  |
| Age, y |  |
| Median (IQR) | 65 (57-73) |
| >65 | 41 (49) |
| Sex |  |
| Male | 60 (72) |
| Female | 23 (28) |
| Weight, kg |  |
| Median (IQR) | 82 (75-95) |
| BMI |  |
| Median (IQR) | 26.8 (24.9 – 30.5) |
| **Reported comorbidities\*** |  |
| Serious heart condition | 15 (18) |
| Chronic lung disease or<br>moderate to severe asthma | 6 (7)<br>9 (11) |
| Immunocompromised | 0 (0) |
| Obese |  |
| BMI > 30 | 23 (29) |
| BMI > 40 | 4 (5) |
| Diabetes | 12 (14) |
| Chronic kidney disease | 2 (2) |
| Liver disease | 0 (0) |
| **Chloroquine treatment** |  |
| Days of therapy |  |
| 1 | 3 (4) |
| 2 | 2 (2) |
| 3 | 6 (7) |
| 4 | 12 (14) |
| 5 | 48 (58) |
| 6 | 9 (11) |
| 7 | 2 (2) |
| 8 | 1 (1) |
| Admitted to |  |
| General ward | 2 (2) |
| Intensive care unit | 81 (98) |
| Maximum number of simultaneous prescriptions<br>of QT-prolonging comedication during<br>chloroquine treatment |  |
| 0 | 23 (28) |
| 1 | 47 (55) |
| 2 | 10 (12) |
| 3 | 3 (4) |
| Drug |  |
| Propofol | 48 (59) |
| Ciprofloxacin | 14 (17) |
| Erythromycin | 13 (16) |
| Amiodarone | 2 (2) |
| Haloperidol | 2 (2) |
| Sotalol | 1 (1) |
| Renal function (CKD-EPI, ml/min) |  |
| Median (IQR) | 73 (52 – 90) |
| Liver |  |
| ASAT >2x ULN | 32 (42% of 75) |
| ALAT >2x ULN | 10 (13% of 75) |
| Outcome |  |
| Deceased | 19 (23) |

\* Comorbidities reported at admission, based on CDC guidance; Groups at higher risk for severe illness

**Figure 1.**
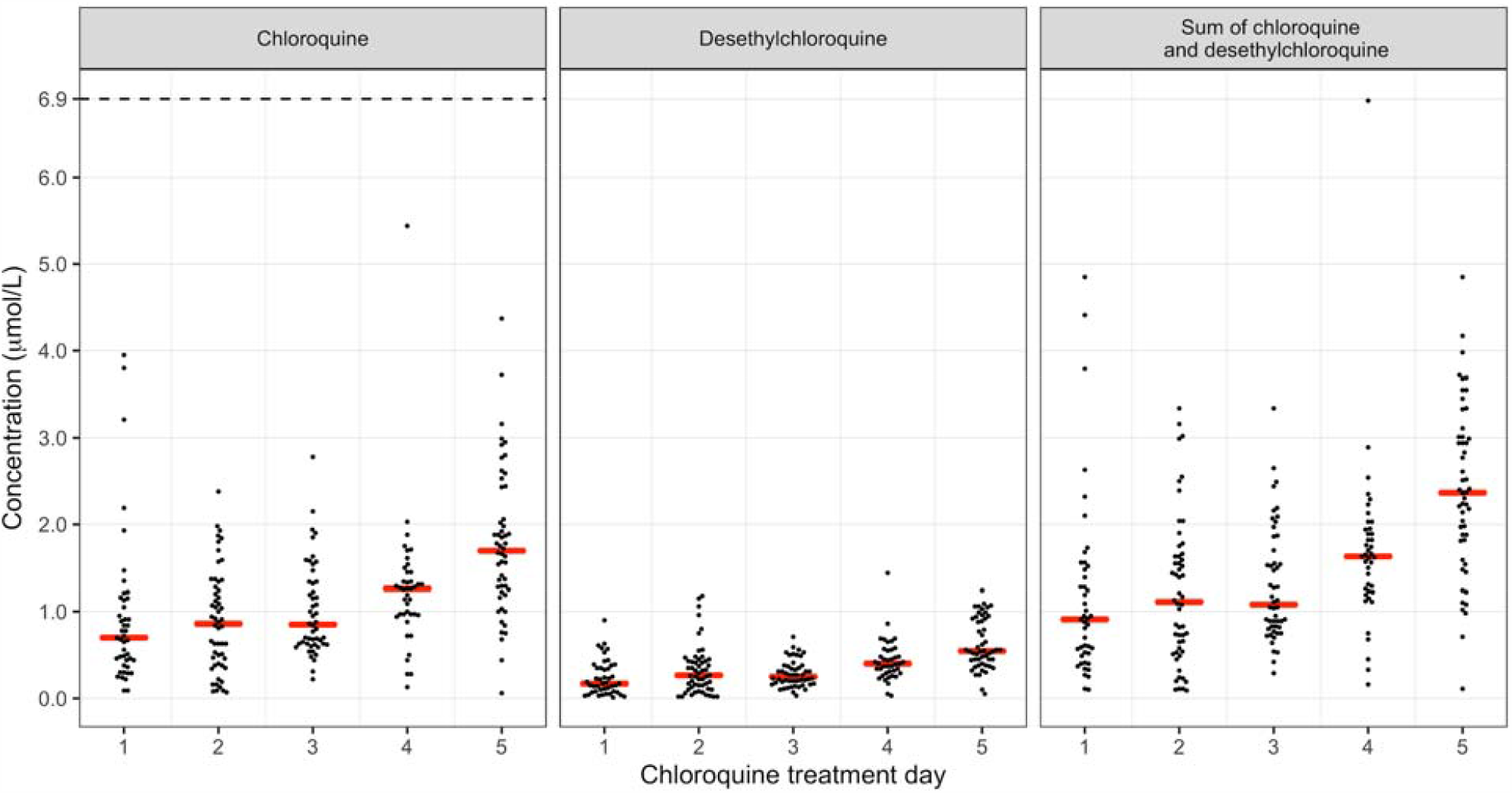
Left panel: Total (free and protein bound) concentration of chloroquine in μmol/L for every treatment day of chloroquine. Middle panel: metabolite desethylchloroquine in μmol/L. Right panel: sum of chloroquine and desethylchloroquine in μmol/L. Dashed line in left panel represents the IC_90_ from the study by Wang et al[2].

QTc measurements at baseline and during chloroquine treatment were available in 41 patients (table 2). In 14 (34%) of these patients a ΔQTc >60 milliseconds was observed after initiation of chloroquine. QTc measurements during chloroquine treatment were available in 69 patients and a QTc ≥500 or ≥600 milliseconds during treatment was observed in 32 (46%) and 7 (10%) of these patients. Torsade de pointes was not reported in any of the patients. No statistical different plasma concentrations of chloroquine were observed in patients with or without QTc >500 ms (p=0.90 with an unpaired t-test on log-transformed pharmacokinetic data).

**Table 2.** QTc time

| Parameter | No (%) of patients (N=83) |
| --- | --- |
| Number of patients with ECG data available | 79 (95) |
| Number of patients with ECG measured at baseline | 51 (61) |
| QTc, Median (IQR) | 450 (425 - 482) |
| >500ms | 7 (14% of 51) |
| Number of patients with ECG measured during treatment | 69 (83) |
| QTc, Median (IQR) | 476 (445 - 501) |
| QTc >500ms at any time during treatment | 32 (46% of 69) |
| QTc >600ms at any time during treatment | 7 (10% of 69) |
| Number of patients with ECG measured at baseline and during treatment | 41 (49%) |
| Maximum QTc prolongation compared to baseline (ms), Median (IQR) | 41 (6-88) |
| QTc prolongation >60ms compared to baseline | 14 (34% of 41) |

## Discussion

The previously *in vitro* determined chloroquine concentrations needed to inhibit viral replication in cell medium were suggested to be clinically achievable[2, 3]. We here show that treatment with chloroquine does not result in plasma concentrations reaching the best-case IC_90_ of 6.9 μM. *In vivo*, chloroquine is metabolized to desethylchloroquine[8]. It is likely that desethylchloroquine has antiviral properties against SARS-CoV-2. Desethylchloroquine is, like chloroquine, a cationic amphiphilic molecule able to accumulate in lysosomes and thereby exerting broad antiviral activity[9]. We found that even the sum of the individual chloroquine and desethylchloroquine concentrations did not exceed the proposed 6.9 μM target (figure 1). Furthermore, chloroquine is known to bind to plasma proteins. In our population, the protein unbound and pharmacologically active fraction of chloroquine and desethylchloroquine in plasma was approximately 20%, while the unbound fraction in cell medium was much higher at approximately 60-80%. Consequently, the gap between the reached pharmacologically active concentrations *in vivo* and *in vitro* IC_90_ is even larger than presented in figure 1 due to this discrepancy. It is known that chloroquine accumulates in lung tissue. The lungs are considered lysosome-rich tissue and the sequestration of cationic amphiphilic drugs like chloroquine in lysosomes, therefore, results in a relatively high overall abundance of chloroquine in lung tissue[10]. However, as the accumulation of chloroquine into the lysosomes is likely to be similar as in other tissues due to the physicochemical properties of chloroquine and the intralysosomal concentration of chloroquine is the putative cause for antiviral effect, it is unlikely that effective exposure is reached in lung tissue.

Patients in our cohort received a total of 3300 mg chloroquine (as chloroquine base) during a 5-day treatment period. Although a simple solution to inadequate exposure would be administration of a higher dose, there are serious concerns for QTc prolongation. Although other drugs than chloroquine and conditions in critically ill patients may cause QTc prolongation, it has been recently shown that high dose chloroquine (a total of 12000 mg as chloroquine base during 10 days) is associated with increased mortality and life-threatening QTc prolongations compared to low dose chloroquine (2700 mg during 5 days) in COVID-19 patients[4]. Further dose increases should, therefore, not be considered. Our findings cast serious doubts on the role of chloroquine for treatment of COVID-19 and underline the necessity of dose finding studies before rushing a drug from the bench to the bedside of a vulnerable population.

## Data Availability

All data are available upon reasonable request to the authors

## Acknowledgement

The authors would like to thank Maaike Sikma, Minou van Seyen, Emilie Gieling, Saskia de Wildt, Soma Bahmany, the Radboudumc intensive care research staff and pharmacy laboratory technicians for facilitating this study.

## References

1. WHO announces COVID-19 outbreak a pandemic [http://www.euro.who.int/en/health-topics/health-emergencies/coronavirus-covid-19/news/news/2020/3/who-announces-covid-19-outbreak-a-pandemic]

2. Wang M, Cao R, Zhang L, Yang X, Liu J, Xu M, Shi Z, Hu Z, Zhong W, Xiao G: Remdesivir and chloroquine effectively inhibit the recently emerged novel coronavirus (2019-nCoV) in vitro. Cell research 2020, 30(3):269–271.

3. Yao X, Ye F, Zhang M, Cui C, Huang B, Niu P, Liu X, Zhao L, Dong E, Song C: In vitro antiviral activity and projection of optimized dosing design of hydroxychloroquine for the treatment of severe acute respiratory syndrome coronavirus 2 (SARS-CoV-2). Clinical Infectious Diseases 2020.

4. Borba MGS, Val FFA, Sampaio VS, Alexandre MAA, Melo GC, Brito M, Mourão MPG, Brito-Sousa JD, Baía-da-Silva D, Guerra MVF: Effect of high vs low doses of chloroquine diphosphate as adjunctive therapy for patients hospitalized with severe acute respiratory syndrome coronavirus 2 (SARS-CoV-2) infection: A randomized clinical trial. JAMA network open 2020, 3(4):e208857–e208857.

5. Fihn SD, Perencevich E, Bradley SM: Caution Needed on the Use of Chloroquine and Hydroxychloroquine for Coronavirus Disease 2019. JAMA network open 2020, 3(4):e209035–e209035.

6. Medicamenteuze behandelopties bij patiënten met COVID-19 (infecties met SARS-CoV-2) [https://swab.nl/nl/covid-19]

7. Drew BJ, Ackerman MJ, Funk M, Gibler WB, Kligfield P, Menon V, Philippides GJ, Roden DM, Zareba W, Cardiology AHAACCCotCoC: Prevention of torsade de pointes in hospital settings: a scientific statement from the American Heart Association and the American College of Cardiology Foundation endorsed by the American Association of Critical-Care Nurses and the International Society for Computerized Electrocardiology. Journal of the American College of Cardiology 2010, 55(9):934–947.

8. Frisk-Holmberg M, Bergqvist Y, Termond E, Domeij-Nyberg B: The single dose kinetics of chloroquine and its major metabolite desethylchloroquine in healthy subjects. European journal of clinical pharmacology 1984, 26(4):521–530.

9. Salata C, Calistri A, Parolin C, Baritussio A, Palù G: Antiviral activity of cationic amphiphilic drugs. Expert review of anti-infective therapy 2017, 15(5):483–492.

10. Assmus F, Houston JB, Galetin A: Incorporation of lysosomal sequestration in the mechanistic model for prediction of tissue distribution of basic drugs. European Journal of Pharmaceutical Sciences 2017, 109:419–430.

